# The Impact of COVID-19 on Care Seeking Behavior of Patients at Tertiary Care Follow-up Clinics: A Cross-Sectional Telephone Survey. Addis Ababa, Ethiopia

**DOI:** 10.1101/2020.11.25.20236224

**Authors:** Tamirat M Aklilu, Workeabeba Abebe, Alemayehu Worku, Henock Tadele, Tewodros Haile, Damte Shimelis, Desalew Mekonen, Wondowossen Amogne, Ayalew Moges, Abebe Habtamu, Rahel Argaw, Sewagegn Yeshiwas, Hyleyesus Adam, Asrat Dimtse, Wakgari Deressa

## Abstract

**Background:** COVID-19, the disease caused by the new coronavirus SARS-CoV-2 is among the most obscure global pandemics resulting in diverse health and economic disruptions. It adversely affects the routine health care delivery and health service uptake by patients. However, its impact on care-seeking behaviour is largely unknown in Ethiopia.

**Objective:** This study was to determine the impact of the pandemic on care-seeking behaviour of patients with chronic health condition at Tikur Anbessa Specialized hospital in Addis Ababa.

**Methods:** A cross-sectional hospital-based survey conducted between May and July 2020 on patients whose appointment was between March to June 2020. Sample of 750 patients were approached using phone call and data collection was done using a pretested questionnaire. After cleaning, the data entered in to IBM SPSS software package for analysis.

**Results:** A total of 644 patients with a median age of 25 years, and M: F ratio of 1:1.01 was described with a response rate of 86%. A loss to follow up, missed medication and death occurred in 70%, 12%, and 1.3% of the patients respectively. In the multivariable logistic regression analysis, patients above 60 years old were more likely to miss follow-up (OR-23.28 (9.32-58.15), P<001). Patients who reported fear of COVID-19 at the hospital were 19 times more likely to miss follow-up (adjusted OR=19.32, 95% CI:10.73-34.79, P<0.001), while patients who reported transportation problems were 6.5 times more likely to miss follow-up (adjusted OR=6.11, 95% CI:3.06-12.17, P<0.001).

**Conclusions:** COVID-19 pandemic affected the care-seeking behaviour of patients with chronic medical condition adversely and the impact was more pronounced among patients with severe disease, fear of COVID19 and with transportation problems. Education on preventive measures of COVID-19, use of phone clinic and improving chronic illness services at the local health institutions may reduce loss to follow-up among these patients.

**What is already known?:**

- As a result of COVID-19, an essential maternal, newborn and child health (MNCH) services in Addis Ababa city showed that first antenatal attendance and under-five pneumonia treatment decreased by 12 and 35%.
- A drop in client flow was ascribed to fear of acquiring COVID-19 at health facilities, limited access due to movement restrictions, and dedication of health facilities as COVID-19 treatment centers.

**What are the new findings?:**

- A cross-sectional hospital-based telephone survey indicated that a loss to follow up, missed medication and death occurred in 70%, 12%, and 1.3% of patients with chronic medical conditions respectively.

**What do the new findings imply?:**

- Fear of COVID-19 and transportation problems are the most commonly stated reasons thus, the finding implies that since health care services to patients with chronic medical conditions is concentrated in specialized referral hospitals mostly aggregated in big cities, patients who travel long distance to get the service are at high risk of Loss to follow up.
- Strengthening the chronic care service at a local health institutions, and promoting COVD-19 preventive measures, may help decrease the LTFU and associated complications.

## Introduction

Coronavirus disease19 (COVID-19) has originated first in China Wuhan Hubei province as a health crisis that struck the global community causing dismal repercussions. It begun late December 2020, when China reported a cluster of pneumonia-like diseases in a family setting, SARS-CoV-2 has spread fast all over the world to involve over 200 countries, making itself a pandemic. The World Health Organization (WHO) notified COVID-19 as pandemic on March 12, 2020(1, 2). Ethiopia had the first COVID-19 positive case confirmed on March 13^th^, 2020 (3). At this time, the government declared a state of emergency labeling the pandemic as a national threat and launched overall preventive measures including advising the community to stay at home, practice strict and frequent hand washing and wearing a face mask. The government also restricted the movement of its people from place to place and laid temporary restrictions in market places, restaurants, shops, cinema houses, religious gatherings and other meetings. There was also a temporary restriction on public transport across-regions and cities (4). During this time, the number of hospital visits dropped sharply at least in large hospitals like Tikur Anbessa Specialized Hospital (TASH). For example, the number of patients who visited the pediatric outpatient and chronic care clinic dropped by 30-70% in April.

Evidence from countries with previous epidemics has revealed that people lose trust in the healthcare system and develop a fear of getting the disease in a health facility. Thus, they avoid seeking treatment for their illness or sending a loved one for medical care, the consequences of which may be increased mortality (5).

Factors like distance from a service center were reported to have a significant association with a reduction in the utilization of the health service delivery during influenza pandemic. Similarly, disease severity was reported to have an association with more health-seeking behavior amongst individuals with influenza-like illness (6, 7). Specific factors predicting the loss to follow up (LTFU) and its consequence in the current pandemic at a tertiary care center were not yet reported in the region to the best of our knowledge. Therefore, the study aimed to assess the impact of the COVID-19 pandemic on health care seeking behavior of patients in the chronic care clinics.

## Patients and Methods

### Study design

A facility-based cross-sectional telephone survey conducted from May and July 2020, among patients who had an appointment between Mid-March and Mid-June 2020 at Tikur Anbessa Specialized Hospital (TASH). TASH is the government-funded hospital located in Addis Ababa, which for the last 48 years has served the community as a national referral center. There are several specialty and sub-speciality clinics for the care of chronically ill patients with over half a million patients are visiting annually. The hospital has 700 beds, 1021 doctors (faculties and postgraduate trainees), 379 nurses, 115 other health professionals and 950 permanent and contract administrative staff dedicated to providing health care services (8). The study was conducted with in several speciality clinics mainly in the pediatric and adult chronic care units. The average monthly Pediatric Neurology, Cardiac, Renal, Gastrointestinal, Chest, Infectious disease (PID), High-risk infant, and Rheumatology clinics visits before the pandemic were 620, 510, 129,100, 60, 300, 230, and 18 patients respectively. The respective figure for adult Neurology, Cardiac, Renal, Gastrointestinal (GI), Chest, Infectious disease(ID) and Rheumatology clinics, was 1307,1653, 565,785,521, 1082 and 343 in respective order.

Majority of the patients used to take lifelong medications uninterrupted or discontinued. Qualified Doctors ranging from consultants to resident doctors run these clinics on regular basis. The hospital renders service to patients based on a different payment system including, Free (13.5%), out of pocket payment (78.4%), Insurance system (6.1%), post-payment (1.9%) and subsidized payment system (0.1%) respectively (8). Since the occurrence of COVID-19, TASH has introduced phone clinics as one mitigation strategy to reach patients who might have missed their appointment and to manage those at risk of COVID-19 while they were at home.

### Source population

adult and pediatric patients with chronic illness having regular follow up at TASH.

### Study population

adult and pediatric patients with chronic illness having scheduled follow up between March 15 and mid of June 2020.

Inclusion criteria: patients and caretakers with chronic illness having scheduled follow up appointment during the study period were included.

Exclusion criteria: Patients and caretakers who did not volunteer to participate and the profile of those who were lost to follow up or died before COVID-19 pandemics were excluded.

### Sample size

Sample size was calculated using a single population proportion formula based on a 50% prevalence estimate of the loss to follow-up of patients in the hospital at 95% confidence level, 4% precision and 20% non-response rate. Accordingly, the sample size required for the study was 750 patients.

### Sampling frame

Sample size was distributed proportionally to each clinic based on the caseload of the specific units. Health management information system (either electronic or registration book) were used to prepare both appointed and attended patients separately. By dividing the allocated sample size number to the total number of patients appointed for the study period gave us the sampling interval. First case was selected using simple random number and using the sampling frame we continued counting until the desired sample size was obtained. by identifying the card numbers and mobile phone numbers of the selected cases, participants were approached for their consent. Independent variables includes: - age, sex, address, patient diagnosis, severity of illness, type of treatment, and year of follow up at TASH. These were extracted from the paper-based chart of the patients. Mobile interview was made to get information’s on patient treatment cost, occupation, parental education, monthly income, means of transport, patient choice to attend their follow up, and whether the telephone call was received or not. Dependent variables were LTFU, missed medications, reasons LTFU, worsening of symptoms, alternative measures taken by patients/caretakers, and the role of local pharmacies and local government health institutions.

### Data collection

A one-day training was given for general practitioners and residents on the use of the data collection tool. Thenafter, demographic and clinical data were abstracted from the charts, to be supplimented by mobile interview. The tool on to which data were collected was adopted, piloted and modified on a group of 20 patients before it was used for data collection. The information gathered from this piloted group were not included in the analysis. In this study, LTFU was defined as any number of missed appointments before the beginning of the study and after the notification of COVID-19 in Ethiopia. Similarly, chronic illness was defined as a disease or condition that usually lasts for 3 months or longer.

### Data Analysis

After manually proof read to maintain data quality, data were entered into SPSS software package version 25 IBM, USA. Data were analyzed using frequency, percentile, mean, median, standard deviation, and interquartile range. To evaluate associations between dependent and independent variables cross-tabulation used first, then bivariate and multivariate regression model was used.

We run logistic regression models to check any dependency among the 10 independent variables using the Odds Ratios (ORs) with their 95% Confidence Intervals (CIs) to quantify any association between an outcome variable and possible predictors. Also, differences in proportions were compared for significance using the chi-square test. A *P*-value <0.05 was considered statistically significant.

### Ethical Considerations

Ethical clearance from the Institutional Review Board of the College of Health Sciences, AAU was obtained. In addition, letter of permission was obtained from the hospital director to access the logbook and the information from the chart. Informed consent was obtained from each patient and caregivers for the interview. Data confidentiality was maintained by avoiding patient identifiers like name and patient card number. Data access was secured by password controlled by the principal investigator.

## Result

Out of the 750 samples, 94 of the patients and caretakers were not traced, there was major information gap in 12 of the 656 interviewed then, we remained to analyze 644. The response rate was 86%.

The age ranged between 1 month and 90 years with the median age of participants was 25 years. Close to half (47.5%) of the participants age were in the category of 19-60 years and the male to Female ratio is 1:1.03, while the median age of females is higher than males (31-IQR-38 and 14.5: IQR-44years) (Table 1).

**Table 1:** Demographic characteristics of patients and their caretakers at the chronic care clinic during COVID-19 pandemic, TASH, 2020.

| Characteristics | Male (%) | Female (%) | Total (%) |
| --- | --- | --- | --- |
| **Age in year** |  |  |  |
| 0-5 years | 82(25.9%) | 52(15.9%) | 134(20.8 %) |
| 6-18 years | 85(26.8%) | 67(20.5. %) | 152(23.6%) |
| 19-60years | 114(36%) | 192(58.8%) | 306(47.5%) |
| >60 years | 36(11.4%) | 16(4.9%) | 52(7.9%) |
| Median age(year) | 14.5(IQR-44) | 31(IQR-38) | 25(IQR-41) |
| Median monthly income(birr) | 3000 | 2000 | 3000 |
| **Occupation** |  |  |  |
| Employed | 95(14.8%) | 86(26.3%) | 181(28.1%) |
| Not employed | 160(50.5%) | 214(65.4%) | 374(58.1%) |
| MI | 62(19.6%) | 27(8.3%) | 89(13.8%) |
| **Total** | **317(49.2%)** | **327(50.8%)** | **644(100%)** |
| MI-missed information |  |  |  |

Hemato-oncology, cardiology and diabetic clinics were the three chronic care clinics with higher caseloads compared to other clinics. 448(70%) of the participants were missing one or more appointments and 53(12%) of them also missed their medications. On the other hand, nearly two third 379(59%) of the participants reported that the local pharmacies did not have the necessary medications and medication price was increased in 167(26%) of the cases. Local Government Health Institutions (LGHI) were considered useful only by 105(16%) of the respondents amid the pandemic. Worsening of symptoms in 68(10.8%) of the case and 8(1.3%) died during the study period (Table 2).

**Table 2:** Clinical characteristics of patients and their caretakers at the chronic care clinic during COVID-19 pandemic, TASH, 2020

| Variables | Males | Females | Total |
| --- | --- | --- | --- |
| Chronic care clinics |  |  |  |
| Cardiac cases | 67(21.1%) | 68(20.8%) | 135(21%) |
| Endocrine cases | 23(7.5%) | 58(17.3%) | 81(12.6%) |
| Gastrointestinal cases | 8(25.2%) | 5(1.5%) | 13(2.0%) |
| Infectious disease cases | 25(7.9%) | 47(14.4%) | 72(11.2%) |
| Neonatal cases | 9(28.4%) | 7(2.1%) | 16(2.5%) |
| Missed appointments | 219(48.9%) | 229(51.1%) | 448(69.6%) |
| Missed medication | 25(47.2%) | 30(52.8%) | 53 (11.8%) |
| Medication obtained at local pharmacy with |  |  |  |
| Increased price |  |  | 167(25.9%) |
| Reduced price |  |  | 2(0.4%) |
| Same price as before |  |  | 71(11.0%) |
| Free |  |  | 26(4.0%) |
| MI |  |  | 378(58.7%) |
| LGHI were helpful during the pandemic |  |  | 105(16.3%) |
| MI |  |  | 70(10.9%) |
| Undesired outcome |  |  |  |
| Worsening of illness | 31(9.8) | 37(11.3%) | 68(10.8%) |
| Mobile phone service |  |  |  |
| Reschedule and link to local HI | 61(9.5%) | 70(10.9%) | 131(20.3%) |
| Refill and dose adjustment | 39(6.1%) | 41(6.4%) | 80(12.4%) |

MI-missed information, LGHI-local government health institute

**Table 3** shows the relationship between independent and the outcome factors in both the univariate and multivariable logistic regression analyses. In the bivariate logistic regression analysis, older patients had higher odds to be lost-to-follow-up compared with the younger patients. Similarly, patients with moderate to severe diagnoses had higher odds to miss the follow-up. Fear of COVID-19 (OR=7.76, 95% CI:5.27-11.41, *P<*0.001), pocket or insurance payment for treatment (OR=1.90, 95% CI:1.34-2.70, *P*<0.001), and transportation problem (OR=2.66, 95% CI: 1.74-4.07, *P*<0.001) were the main factors associated with lost-to-follow-up of patients.

**Table 3.**
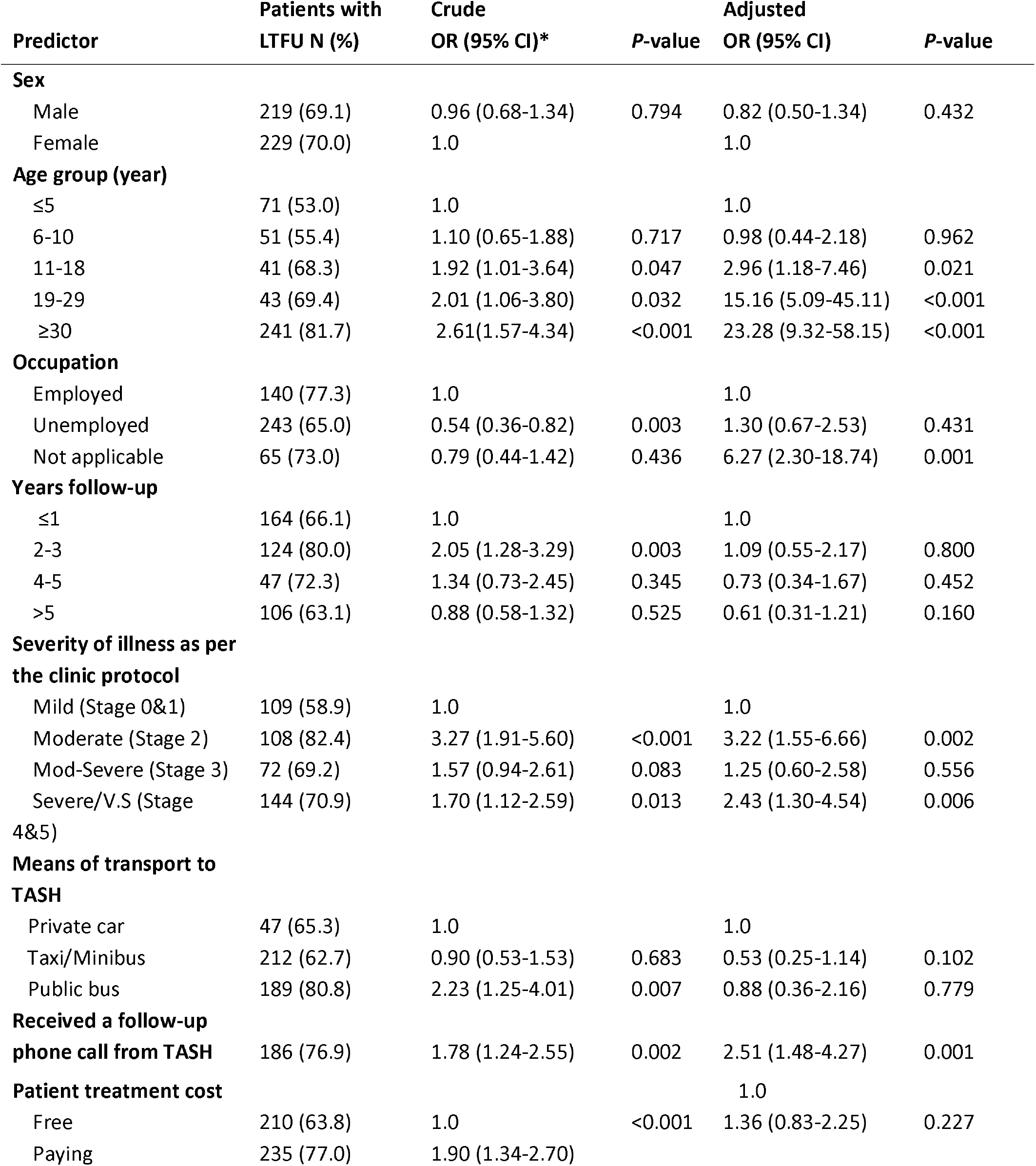
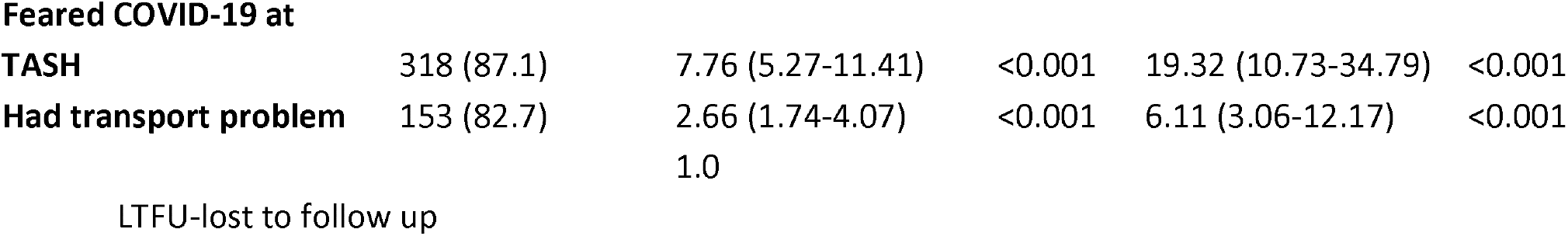
Factors associated with loss-to-follow-up of patients at a tertiary treatment center in the bivariate and multivariable analyses, TASH, 2020

In the multivariable logistic regression analysis model, (Table 3) performed using the 10 independent variables, older patients again had higher odds to miss a follow-up. The odds of loss to follow up among patients who reported the fear of COVID-19 were 19 times higher than among patients who did not have fear. (adjusted OR=19.32, 95% CI:10.73-34.79, *P*<0.001), the odds of loss to follow up among patients who reported transportation problems were 6.5 higher than among those who don’t have transport problem (adjusted OR=6.11, 95% CI:3.06-12.17, *P<*0.001). Factors such as sex, years of follow-up, distance and means of transport, and treatment costs did not have a significant influence on the loss-to-follow-up. The multivariable binary logistic regression model presented in Table 3 had goodness-of-fit under the Hosmer-Lemeshow test (X2=13.32, *P*=0.101), and the full model containing all predictors was statistically significant X2 (8, N=604) = 292.6, *P*<0.001. The model as a whole explained between 38% (Cox-Snell R2) and 54% (Nagelkerke R2) of the variance in satisfaction level. There were eight mortality cases, cancer being the diagnosis in six of them, diabetes mellitus in one of them, and valvular heart disease in the remaining one. All the mortality cases were lost to follow up during the pandemic while; half of them also missed their medications. In all of them, disease severity was moderate to severe. The reasons for missed appointment were fear of COVID-19 infection in 6 of them. The majority lived in an area with more than 200 km away from the referral hospital (TASH).

As depicted in figure 2, Fear of COVID-19 infection, and lack of transport were the main reasons to be lost for follow up. Other COVID related reasons like family member illness by COVID-19 were also shown.

**Figure 1.**
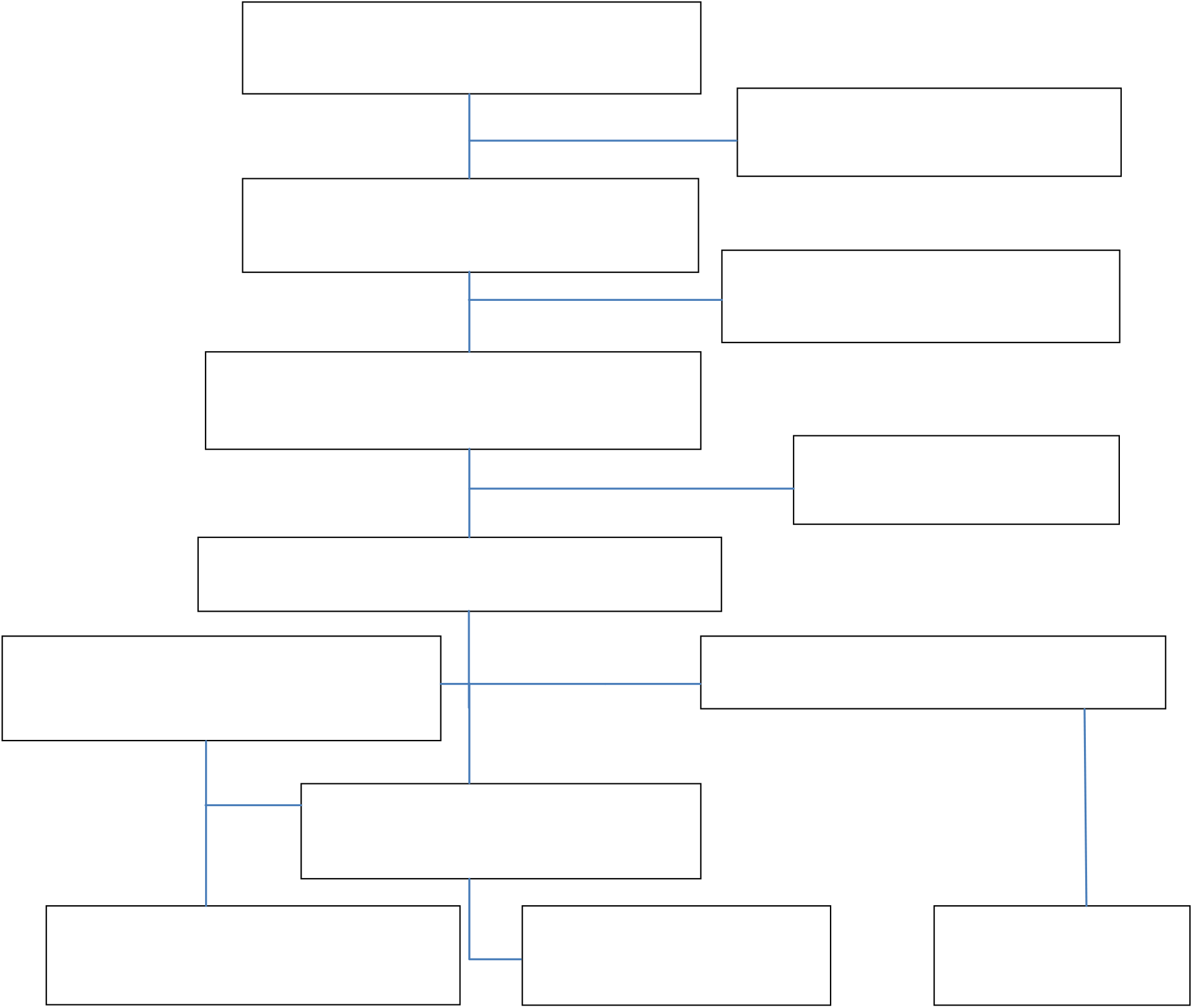
Participant flowchart included in the study, TASH, 2020

**Figure 2:**
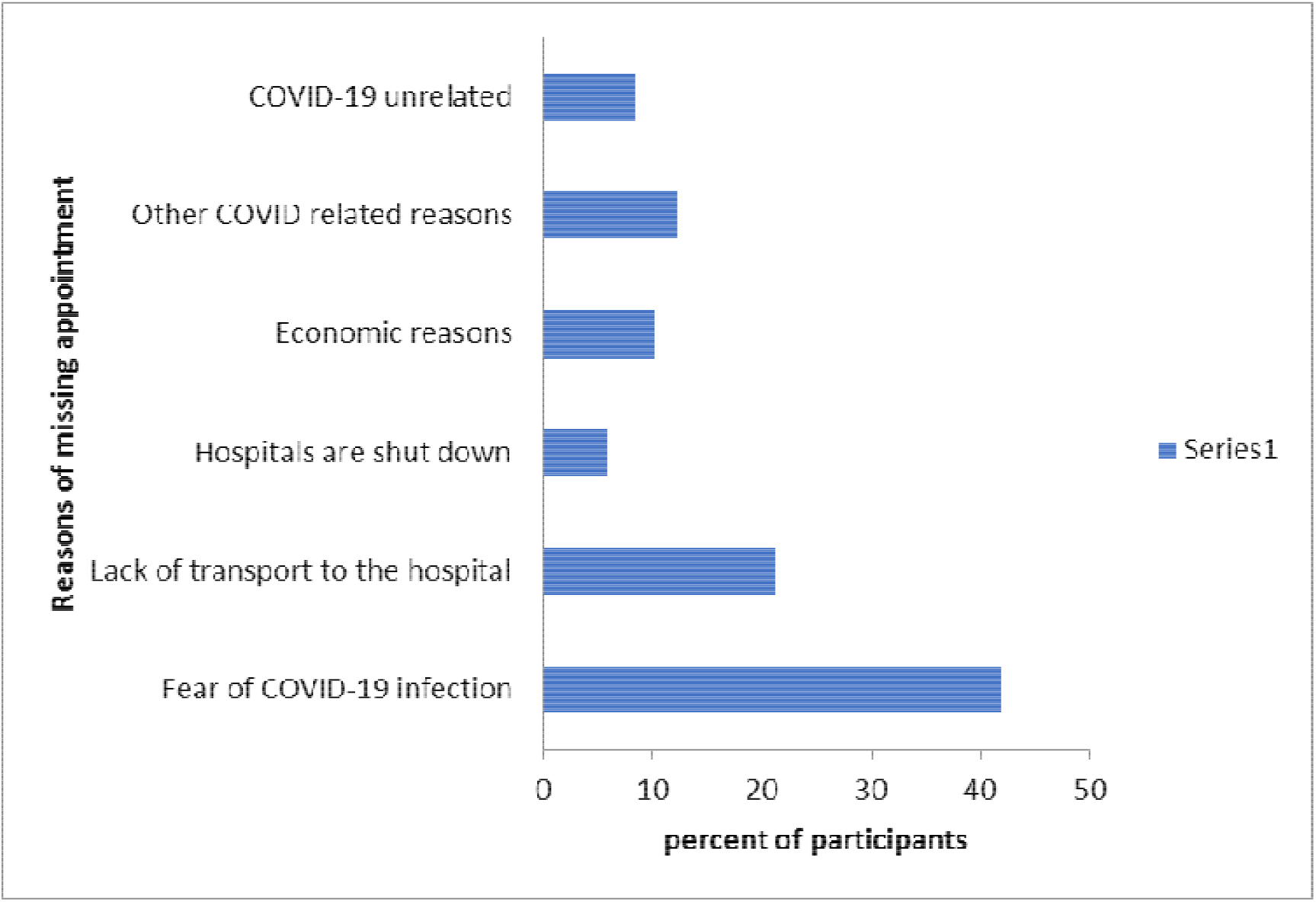
Reasons for missing hospital appointment as responded by participants TASH, 2020.

Figure 2 showed types of medications that patients were taking on while having follow up at TASH. Multiple medications, antiepileptics, chemotherapies and antiretroviral drugs were among the common ones. Majority of the participants did not seek alternative ways to compensate for their missed appointments. However, self-refill is the most common alternative action taken by the patients figure 3.

**Figure 3:**
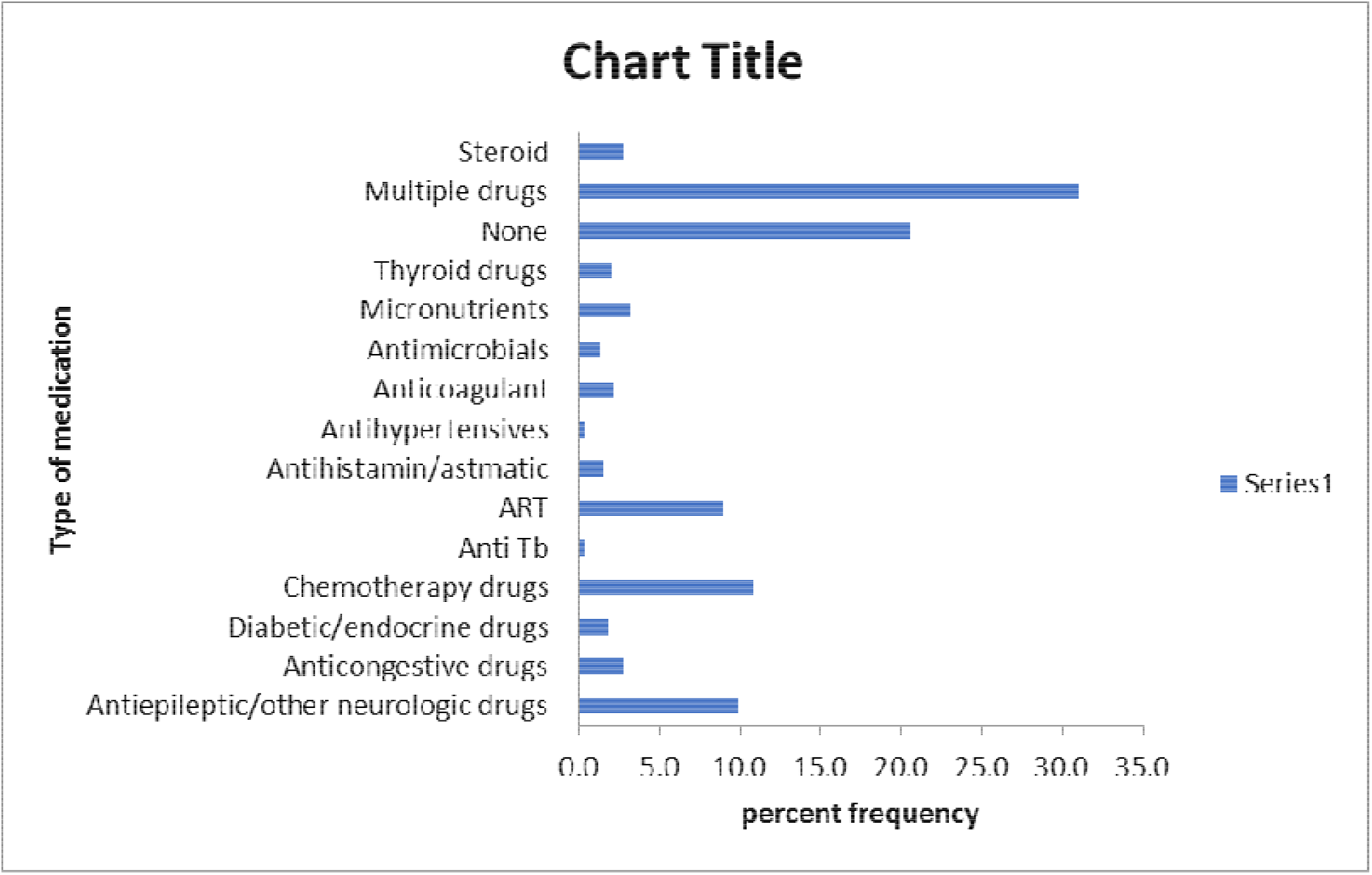
Types of medications patients were on while on follow up at TASH, 2020.

As depicted in Figure 4, 207(27.6%) of the patients responded to the question about alternative source of medication when they were missing the appointment. Of these 132(20.5%) did not search for any alternative source of treatment. 29(4.5%) of them bought their medications without prescription. The rest sought health care provider (HCP) in the nearby private or government health institutions. 11 (1.7%) of them opted for Holy water.

**Figure 4:**
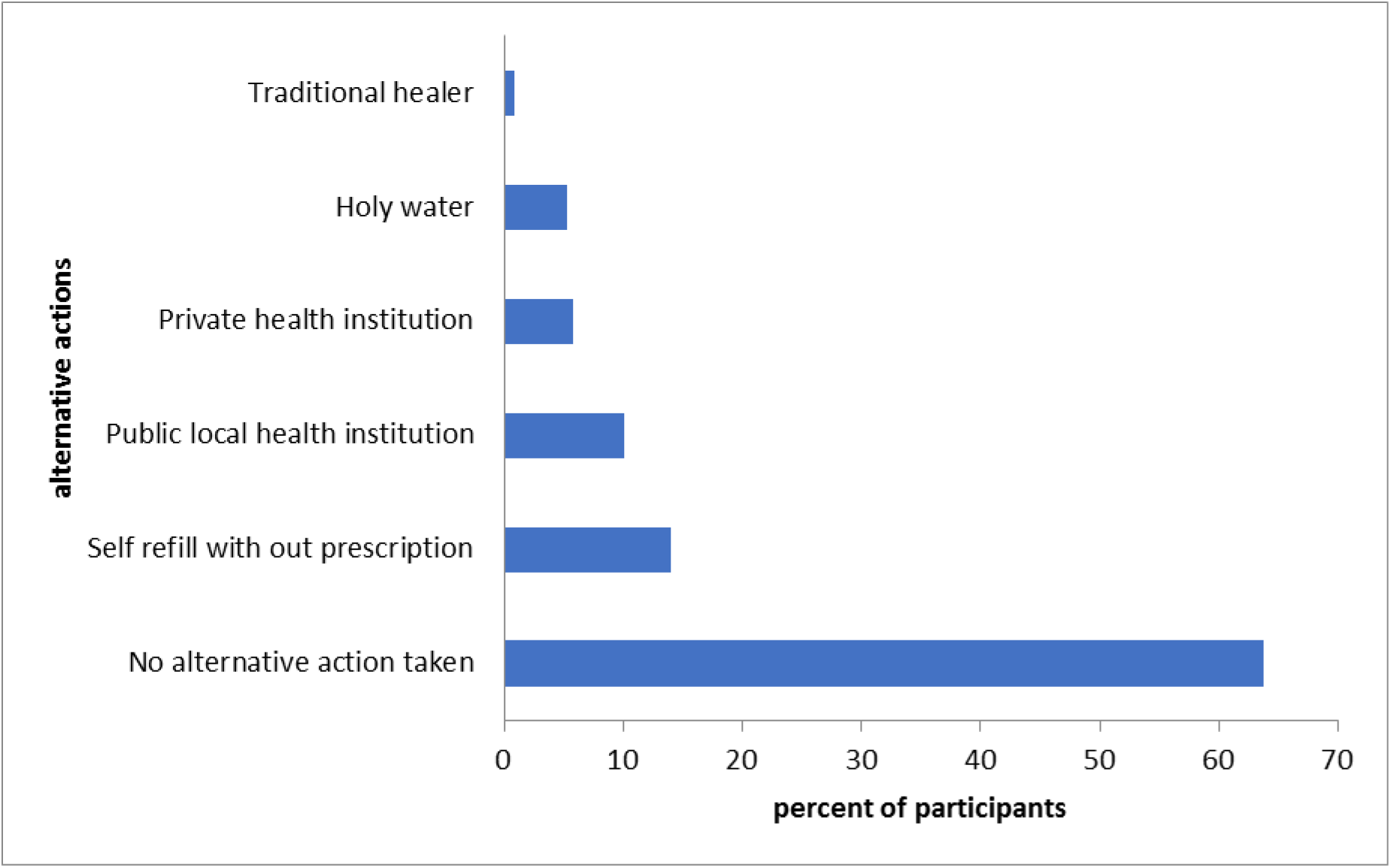
patients /caretakers took alternative actions as they missed their hospital appointments at TASH 2020.

## Discussion

The study demonstrated that there was a decreased in the care seeking behavior of patients at follow-up clinic of TASH in the first 3 months of COVID-19 pandemic, March 15 - June 15, 2020. This was more pronounced in older age and moderate to severely ill patients.

Patients and caregivers who have had received phone calls from the hospital prior to the study also missed follow up. In addition, patients who reported fear of acquiring COVID-19 infection in the hospital and transportation problem as the main reason of absence from the clinics were among those who missed the follow-up appointment more frequently than their counterparts did. The finding that 70% of the participants missing appointment during March 15 to June 15, 2020, were comparable to the experience of others during former epidemics. A similar response to the Ebola epidemic in 2013/14 has also been observed in Sera Leon, Guinea and Liberia ((9, 10). For example, in Sierra Leone, two studies including primary health facilities and facilities offering basic emergency obstetric care observed that facility-based deliveries were reduced by 27–37%. In Guinea, a sharp decline of 74–81% in delivery care seen in two of the regions most affected by Ebola virus disease. On the other hand, essential health service disruption in Pakistan by the COVID-19 pandemic was estimated to increase by 22 per cent and as many as 5,424,900 fewer children would have received oral antibiotics for pneumonia, 5,441,800 fewer would have received DPT vaccinations (11, 12).

Although the WHO reported a 4% LTFU rate globally, these data are at odds with experience in the field and there is growing concern over the accuracy of population-level estimates (13). LTFU from various health care services were common in the population with socio-economic difficulties even before COVID-19 pandemic.

Reports from various LMICs revealed that for patients with hypertension, diabetes, and epilepsy, the LTFU rate was 22% to 42%, 35%, and 27% to 34% respectively (14, 15). Similarly, literature from child-nutrition programs and women after screening positive for cervical cancer, the LTFU rate showed a similar picture (16, 17). The 70% LTFU rate in the current study may be expected in low-income community particularly in the face of COVID-19 pandemics.

The high LTFU rate observed in the current study can be explained by the temporary lockdown and abandoning of elective surgeries as some of the preventive measures to halt the spread of the virus. Earlier report in this country amid the pandemic, showed a decreased in attendance of essential maternal, newborn, child health services in Addis Ababa city and under-five pneumonia treatment by 12% and 35% respectively(18).

These reasons were in line of evidences from many countries. Cancellations of planned treatments, a decrease in public transport available and a lack of staff because health workers were reassigned to support COVID19 services were among the few. On the other hand, in one in five reporting countries, the main reasons for discontinuing services were a shortage of medicines, diagnostics and other technologies (19). Similar chronic illness care disruptions in Romania, both in the general and specialist medical services were significantly limited because of the rules imposed to prevent COVID-19 propagation (20).

Old age was an important determinant factor to the LTFU in the current study. Report from COVID-19 hard hit countries; mortality and severe disease were higher among aged people (18-21). Therefore, it is highly probable that older patients avoided coming to the hospital may be due to consideration of their vulnerability. Fear of Covid-19 was found to be a very strong predictor of LTFU in the current study. Similar observation made elsewhere in this country, north-west Ethiopia, acquiring COVID-19 virus was highly threatening for nearly half (52.54%) of patients with chronic diseases (19, 20).

In this study, those who reported transportation problem missed their follow up visit more frequently than their counterparts did. It was observed that the novel coronavirus has had a profound, dramatic impact on the transport system. Because of the contagion, risk caused by the COVID 19 leads drastic reduction in transport services. It was suggested that limiting transport for health reason was there in the past, but the scale of the restrictions in the current pandemic was unprecedented (21). On the other hand, it was an undeniable fact that transport and mobility have become a potential site for the spread of the virus—especially in situations where transport is provisioned with enclosed spaces and large concentrations of people, as is often the case in poor communities like ours (22, 23). Contrary to the developed world where, many aspects of daily lives have moved online, in low-income countries it is an abrogate to move from one place to another to get service. Preventive measures like restricting transportation with in a country thus, affects patients and caretakers directly affecting those who necessarily taking uninterrupted medications (21, 24).

Seventy-three per cent of the respondents in the current study did not consider local government health institutions useful during the pandemic. According to WHO Essential NCD package, making chronic illness care, available at primary care centers, will help to relieve the stress of going far distance to tertiary centers particularly during COVID-19 pandemic (21).

In fact, the current finding is contradictory to the Ethiopian, Health Service Transformation Plan 2015/2016 that reported 23% to 76 % of local government health institutions were offering services to people with NCD (25).

In this study, over three-quarters of the cases have received a telephone call. The high figure in LTFU should have been also the result of the mobile phone call made by the hospital. During the call, patients were rescheduled, told to buy their medications at local pharmacies or few requested to come to the hospital for further evaluation (Table 2). As the mobile phone call was delivered by the hospital in an attempt to link the patients during this difficult time, it did not follow a uniform standard and the outcome is difficult to evaluate. In fact, it may not be considered as an important determinant factor for loss to follow up. However, since there was no standardized protocol and it may not be accessible to all patients, mobile phone service may be false reassurances to patients to miss their hospital follow up, the current finding may be important to influence LTFU negatively.

It was argued that in the absence of clear protocols and standardized guidelines for the delivery of telemedicine consultations, telemedicine might result in harm than benefit. The implementation in the field of such alternative consultation requires the development of relevant standards and procedures (26). On the other hand, it was also emphasized that twining the mobile phone clinic (telemedicine) with tele pharmacy is important to increase healthcare delivery while decreasing the SARS COV-2 spread (27-29).

A quarter (26%) participants responded that medication price has increased following COVID-19 confirmation. Similar observations were made in other reports as well. Furthermore, WHO stated that COVID-19 pandemic has affected the availability of medicines in many countries (26, 27). National, as well as an international transportation problem, the workforce and the lockdown measures, have been among the few reasons cited as contributing factors (27, 28). Global Health Action reported that medicines for non-communicable diseases in many countries were not available. Countries are already facing the problems when things were made worse by the COVID-19 pandemic (30)

The other observation made in this study was patients with moderate to severe medical conditions were loss to follow up more frequently. A contrary observation was made in the European study, among influenza patients where disease severity was reported to have an association with more health-seeking behavior (6). Experience also told that, the severity of COVID-19 infection is directly related to severity of the underlying medical conditions (30). However, the two scenarios are too different as the former is about risk exposure to viral disease (COVID-19) while the latter is getting sick by the virus (influenza) itself.

In the current study, 11.8% of the loss to follow up cases also missed their medications. Those who did not miss medications despite LTFU, have given information on further inquiries, that they had extra pills at home, or they were able to buy their medications with old prescriptions. A similar observation was reported during Ebola epidemics (31).

As depicted in Figure 4, Patients had opted to the different alternative medicine. Apart from the current pandemic, report showed that the use of alternative medicines are commonly practiced among patients with hypertension and other NCDs; faith in treatment with alternative medicine, dissatisfaction with conventional medicine and the desire to reduce medication costs are the reasons for opting(32).

The use of undocumented prescription and its consequence has been discussed (33). Self-medication is an intermittent or continued use of prescribed drugs for chronic or recurrent disease or symptoms; a major shortfall of it is the lack of clinical evaluation of the condition by a trained medical professional, which could result in missed diagnoses and delays in appropriate treatments (34).

In this study, we received report of 8 deaths between the beginning of Covid 19 pandemics and the end of the study among the study group. Whether the death was directly related to COVID-19 or not was not known. However, their disease severity and the probable shortage of the medicines due to the lockdown and the psychosocial impact of COVID-19 may have contributed to their death. The fact that most of them were a cancer case would mean a high risk during crises time like COVID-19.

### Limitation of the study

This is a telephone surveys where many were not accessible. The inaccessibility may be due to nonfunctioning cellphones, battery death because of frequent power interruption, no smartphones and low or non-response is also common. Further, the fact that one-third of the patients received a telephone call from the hospital would dilute our observation as three-quarters of them missed their follow appointment.

## Conclusion

COVID-19 pandemic impacted the care-seeking behavior of patients with chronic health condition negatively and the impact was more severe among patients with severe disease, those with fear of COVID-19, and transportation problem. Education on preventive measures of COVID-19 and improving chronic illness care services by the local health institutions including availability of essential drugs may reduce the dire effect of the loss to follow up among patients with a chronic medical condition. The practice of phone service in LMIC, should be based on the local standard operation procedure taking into account the communities’ interest and access to technology.

## Data Availability

All relevant data are included in the manuscript.

## Authors affiliations

1. Department of Pediatrics and Child Health, College of Health Sciences, Addis Ababa University
2. School of Public Health, College of Health Sciences, Addis Ababa University
3. Department of Internal Medicine, College of Health Sciences, Addis Ababa University

## Acknowledgements

We thank the following individuals who tirelessly worked in collating the data-Dr. Ruth Yizengaw, Dr Fikreab Bisrat, Dr Abel Tenaw, Dr Tigist Abebaw,Dr Nathan Wondwossen, Dr Yibeltal Amsalu, Sr Tiruwork Tadesse, Dr Yodit Abreham, Dr Elias Simenew and Dr Frezer Milashu. We thank the nursing staff at the cardiac, neurology, renal, diabetic, GI, ID, Chest, High Risk, Hemato-oncology and rheumatology clinics(both adult and pediatric) for collaborating with the data collectors.We also thank W/o Teshay and W/t Hareg from department of Pediatrics who helped in assisting the administrative works. Our thank extends to W/o Elsa and the card room staff for their kind cooperation with chart retrieval. We are also grateful to Dr Nicola Ayers for her contribution in the language editing.

Last but not least, we thank the patients and their caretakers for allowing us to conduct the study using their profile.

## Contributors

TMA incepted the research question and developed the design, the proposal,and the write-up. WAA-reviewed the proposal and involved in the write-up. AW revised the statistical analysis and involved in the writ up. WD reviewed the proposal, and did the statistical analysis. WD also involved in the write-up. TH, DM, DS, HT and WA all involved in the review of the proposal and the write-up. AM, AH, RA, SY, HA, AD collaborated in data acquisition. They also reviewed the manuscript.

## Patient and public involvement

Patient and public were not involved in the design or conduct or reporting or dissemination plans of the research.

## Patient consent for publication

Obtained.

## Ethics approval

AAU college of Health Sciences Institutional review board appropriated the study(Protocol no. 042/20/SPH).

## Provenance and peer review

Editorial commissioned; externally peer reviewed.

## Data availability statement

All data relevant to the study are included in the report.

## Open access

This is an open access article distributed in accordance with the Creative Commons Attribution Non Commercial (CC BY-NC 4.0) license, which permits others to distribute, remix, adapt, build upon this work non-commercially, and license their derivative works on different terms, provided the original work is properly cited, appropriate credit is given, any changes made indicated, and the use is non-commercial. See: http://creativecommons.org/licenses/by-nc/4.0/.

